# Pediatric Gastrointestinal Outcomes During the Post-Acute Phase of COVID-19: Findings from RECOVER Initiative from 29 Hospitals in the US

**DOI:** 10.1101/2024.05.21.24307699

**Authors:** Dazheng Zhang, Ronen Stein, Yiwen Lu, Ting Zhou, Yuqing Lei, Lu Li, Jiajie Chen, Jonathan Arnold, Michael J. Becich, Elizabeth A. Chrischilles, Cynthia H. Chuang, Dimitri A Christakis, Daniel Fort, Carol R. Geary, Mady Hornig, Rainu Kaushal, David M. Liebovitz, Abu Saleh Mohammad Mosa, Hiroki Morizono, Parsa Mirhaji, Jennifer L. Dotson, Claudia Pulgarin, Marion R. Sills, Srinivasan Suresh, David A. Williams, Robert N. Baldassano, Christopher B. Forrest, Yong Chen, the RECOVER Initiative

## Abstract

**Importance:** The profile of gastrointestinal (GI) outcomes that may affect children in post-acute and chronic phases of COVID-19 remains unclear.

**Objective:** To investigate the risks of GI symptoms and disorders during the post-acute phase (28 days to 179 days after SARS-CoV-2 infection) and the chronic phase (180 days to 729 days after SARS-CoV-2 infection) in the pediatric population.

**Design:** We used a retrospective cohort design from March 2020 to Sept 2023.

**Setting:** twenty-nine healthcare institutions.

**Participants:** A total of 413,455 patients aged not above 18 with SARS-CoV-2 infection and 1,163,478 patients without SARS-CoV-2 infection.

**Exposures:** Documented SARS-CoV-2 infection, including positive polymerase chain reaction (PCR), serology, or antigen tests for SARS-CoV-2, or diagnoses of COVID-19 and COVID-related conditions.

**Main Outcome(s) and Measure(s):** Prespecified GI symptoms and disorders during two intervals: post-acute phase and chronic phase following the documented SARS-CoV-2 infection. The adjusted risk ratio (aRR) was determined using a stratified Poisson regression model, with strata computed based on the propensity score.

**Results:** Our cohort comprised 1,576,933 patients, with females representing 48.0% of the sample. The analysis revealed that children with SARS-CoV-2 infection had an increased risk of developing at least one GI symptom or disorder in both the post-acute (8.64% vs. 6.85%; aRR 1.25, 95% CI 1.24-1.27) and chronic phases (12.60% vs. 9.47%; aRR 1.28, 95% CI 1.26-1.30) compared to uninfected peers. Specifically, the risk of abdominal pain was higher in COVID-19 positive patients during the post-acute phase (2.54% vs. 2.06%; aRR 1.14, 95% CI 1.11-1.17) and chronic phase (4.57% vs. 3.40%; aRR 1.24, 95% CI 1.22-1.27).

**Conclusions and Relevance:** In the post-acute phase or chronic phase of COVID-19, the risk of GI symptoms and disorders was increased for COVID-positive patients in the pediatric population.

**Key Points:** *Question:* Does COVID-19 increase the risk of gastrointestinal (GI) symptoms and diseases during the post-acute phase in children and adolescents?

*Findings:* Newly diagnosed GI symptoms and disorders such as diarrhea, constipation, and vomiting are seen more commonly in children and adolescents with SARS-CoV-2 infection.

*Meaning:* Clinicians need to be mindful that after SARS-CoV-2 infection in children, lingering GI symptoms without a unifying diagnosis may be more common than among uninfected children.

## Introduction

Long COVID ^1–11^, also known as the Post-Acute Sequelae of SARS-CoV-2 (PASC), was initially identified primarily in adults, but its emergence has also raised concerns for the pediatric population^12–14^. In the United States (US), depending on the definition used and the study population, PASC is reported to affect 1.3% of children, a rate lower than that reported in some studies for adults (6.7%)^15,16^. This may not be surprising as there are important differences in the presentation and outcomes of acute SARS-CoV-2 infection between children and adults. Children can have different symptoms compared to adults and tend to have a milder disease course, or even one that is asymptomatic, with a lower risk of hospitalization or death– particularly children without pre-existing conditions^17–19^. Given these differences in acute infection, as well as the differences in prevalence between children and adults, the characteristics of PASC require further study in children.

Multiple organ systems can be affected in PASC, including the gastrointestinal (GI) tract^20–25^. The risk of GI symptoms and disorders, including abdominal pain, diarrhea, constipation, vomiting, bloating, irritable bowel syndrome (IBS), and gastroesophageal reflux disease (GERD) was increased in the one year after SARS-CoV-2 infection amongst adults.^10^ This phenomenon of chronic GI symptoms potentially developing after infection was well-described in adults before the SARS-CoV-2 pandemic with post-infectious IBS associated with acute gastroenteritis^26,27^ and post-viral gastroparesis seen in association with several respiratory and GI viruses^28^. Children are also at risk for developing an array of lingering GI symptoms after an acute infection but in contrast to adults, pediatric patients predominantly present with chronic abdominal pain and constipation^29^. As the data regarding the long-term effect of SARS-CoV-2 on the GI tract amongst children is limited^30^, it is unclear if children have the same risks of GI conditions during the post-acute phase of COVID-19 as has been seen with adults.

In this retrospective cohort study, we aimed to investigate the risk of GI symptoms and disorders during the post-acute and chronic phases of COVID-19 illness among children and adolescents from twenty-nine children’s hospitals and health institutions in the US. Specifically, we first estimated the incidence of any GI symptoms and disorders associated with SARS-CoV-2 infection and evaluated whether the SARS-CoV-2 infection increases the risk of GI symptoms and disorders. To the best of our knowledge, our investigation has the longest follow-up duration (two years) and is the largest study reported for children and adolescents that assesses the risks of COVID-19 infection for GI symptoms and diseases in the US.

## Methods

### Data Sources

This study is part of the NIH Researching COVID to Enhance Recovery (RECOVER) Initiative (https://recovercovid.org/), which aims to learn the long-term effects of COVID-19. The twenty-nine health institutions include Children’s Hospital of Philadelphia (CHOP), Cincinnati Children’s Hospital Medical Center, Children’s Hospital Colorado, Columbia University Irving Medical Center, Duke University, The Ohio State University, Ann & Robert H. Lurie Children’s Hospital of Chicago, Medical College of Wisconsin, University of Michigan, University of Missouri, Montefiore, Medical University of South Carolina, Children’s National Medical Center, Nationwide Children’s Hospital, Nicklaus Children’s Hospital, the University of Nebraska Medical Center, The Nemours Foundation, Northwestern University, New York University School of Medicine, OCHIN, Inc., Ochsner Health System, University of Pittsburgh, Penn State University, Seattle Children’s Hospital, Stanford University, University of California San Francisco, University of South Florida and Tampa, Vanderbilt University Medical Center, and Weill Cornell Medical College. The data are standardized according to the National Patient-centered Clinical Research Network^31^ Common Data Model.

### Cohort Construction

Our study period spanned from March 2020 to September 2023, with a cohort entry date cutoff of March 2023 (Each participant had at least six-month follow-up period). Our study focused on individuals aged not above 18 who had at least one visit within 24 months to 7 days before the index date and subsequently had another encounter 28 days to 729 days after the index date. For the cohort defined as having a positive COVID-19 status, we included patients who had at least one documented SARS-CoV-2 infection, including positive polymerase chain reaction (PCR) tests, serology, antigen tests, diagnosed COVID-19, or PASC. We excluded the patients with the diagnosis of multisystem inflammatory syndrome in children (MIS-C) from the COVID-19 positive cohort. The detailed exclusion criteria were summarized in **Figure 1**. The cohort entry date was defined as the first index date of the documented SARS-CoV-2 infection. As for the COVID-19 negative cohort, we selected pediatric patients who did not have the documented SARS-CoV-2 infection or MIS-C diagnosis during the same study period and who had at least one negative COVID-19 test result. The index date for the cohort with COVID-19 negative was determined by randomly choosing a negative test result.

**Figure 1.**
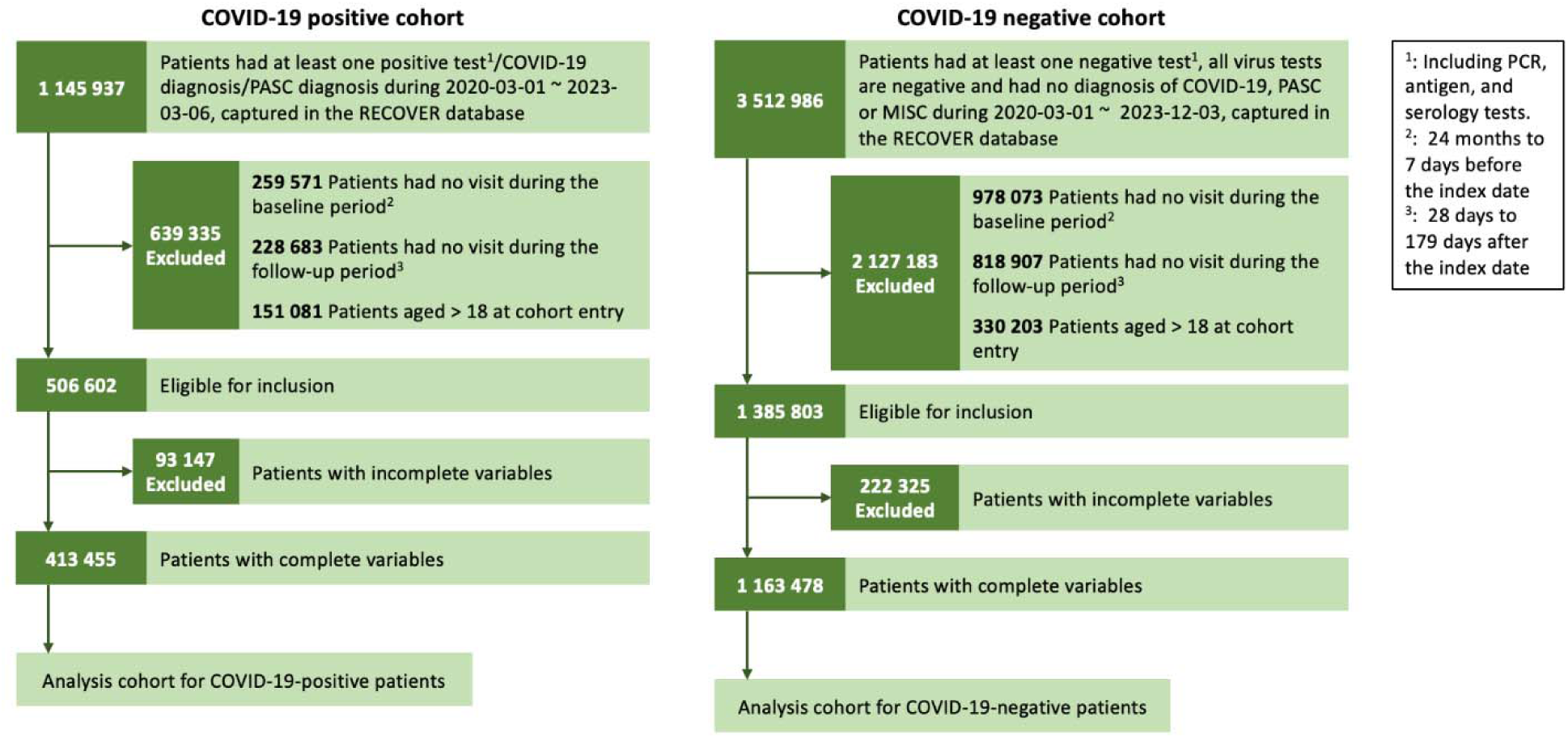
Attrition table for COVID-19 positive cohort and COVID-19 negative cohort.

### Defining GI Outcomes

We explored nine predefined GI symptoms and disorders within each follow-up period (28 days to 729 days after the cohort entry date). The GI signs or symptoms were abdominal pain, bloating, constipation, diarrhea, nausea, and vomiting. The GI disorders were functional dyspepsia, GERD, and IBS. The GI symptoms and disorders were systematically categorized using the ICD-10-CM codes provided in **Section S13** in the Supplement.

### Patient Characteristics

The baseline covariates included factors such as age at cohort entry, sex (female, male), race and ethnicity (Non-Hispanic White (NHW), Non-Hispanic Black (NHB), Hispanic, Asian American/Pacific Islander (AAPI), Multiple, Other/Unknown), obesity status during the baseline period (from 24 months to 7 days before the cohort entry date). We also considered the month of cohort entry from March 2020 to March 2023, chronic condition status as defined by the Pediatric Medical Complexity Algorithm^32,33^ (PMCA)—which categorizes conditions into no chronic condition, non-complex chronic condition, and complex chronic condition. Additionally, we included the site index for the cohort entry, and healthcare utilization indicated by the number of the inpatient, outpatient, and emergency department visits during the baseline period. We analyzed the number of unique medications or prescriptions (categorized as 0, 1, 2, >=3) and the count of negative tests conducted from 24 months to 7 days before the cohort entry date, also grouped as 0, 1, 2, >=3.

### Statistical Methods

To illustrate the incidence of GI symptoms and disorders, we divided the number of patients having GI symptoms or disorders during the follow-up period but not at baseline by the total number of population at risk. We investigated the adjusted risk ratio (aRR) of the prespecified GI symptoms and disorders during the post-acute phase (from 28 days to 179 days after the cohort entry date) and the chronic phase (from 180 days to 729 days after the cohort entry date). To further enhance our analysis, we also investigated the risk of composites of any prespecified GI symptoms, GI disorders, and visits related to GI during the post-acute phase or chronic phase. Before fitting the model, we employed a cutoff incidence value of 0.1%^34^ to prevent the implications of model overfitting for rare GI outcomes. To ensure the covariate balance between the COVID-19 positive and negative cohorts among the patient characteristics served as the confounders listed above, we used propensity-score-based stratification. The propensity score model, derived from a logistic regression model with the outcome as the indicator of COVID-19 positivity, incorporated the patient characteristics as covariates. Confounder assessment utilized standardized mean differences (SMDs), with a cutoff value set at less than 0.1^35^. The aRR estimated risk quantification via a Poisson regression model stratified by the propensity score for the post-acute or chronic phases. Additionally, standard errors were reported based on sandwich estimators^36^ to enhance result robustness.

### Sensitivity Analysis

To ensure the robustness of our findings regarding GI symptoms and disorders, we performed subgroup analysis based on age, race/ethnicity, obesity status, sex, and medical history of diabetes (yes or no) or cardiovascular disorders (yes or no) presented in **Sections S2-S7**. To further investigate the impact of hospitalization during the acute phase of COVID-19, we implemented our analysis method for the ICU admission, hospitalized, and non-hospitalized groups in **Section S8**. Additionally, we explored the influence of varying severity levels of the acute phase (0 to 28 days after the cohort entry date) of COVID-19^37^ by presenting results for the asymptomatic, mild, moderate, and severe groups in **Section S9**. In **Section S10**, we explored the risk of gastrointestinal symptoms and disorders associated with different SARS-CoV-2 virus variants, including pre-Delta, Delta, and Omicron. The ICD-10-CM codes of diabetes and cardiovascular diseases were listed in **Section S14**.

### Negative Control Outcomes

Although we have incorporated an extensive array of confounders, the regression model might still be susceptible to residual bias, including unmeasured confounder bias. To reduce such bias from the estimation of aRR, we employed a calibration approach^38,39^ involving thirty-six negative control outcomes. Details regarding the calibration method were elucidated in **Section S11** of the Supplement, and the full list of negative control outcomes was provided in **Section S12**.

## Results

We presented the descriptive analysis of both the COVID-19 positive and negative cohorts in **Table 1**. Our analysis included 1,576,933 patients. Among them, 756,618 (48.0%) were female. 74,369 (4.7%) were AAPI, 376, 728 (23.9%) were Hispanic, 267,056 (16.9%) were NHB, 671,358 (42.6%) were NHW, and 147,627 (9.4%) were other/unknowns, and 39,795 (2.5%) were multiple races.

**Table 1.** Patient characteristics of COVID-19 negative cohort, COVID-19 positive cohort, and the overall cohort.

|  | Negative<br>(N=1,163,478) | Positive<br>(N=413,455) | Overall<br>(N=1,576,933) |
| --- | --- | --- | --- |
| **Age at Cohort Entry** |  |  |  |
| <1 | 132283 (11.4%) | 57135 (13.8%) | 189418 (12.0%) |
| 1-4 | 349078 (30.0%) | 105599 (25.5%) | 454677 (28.8%) |
| 5-11 | 366863 (31.5%) | 121287 (29.3%) | 488150 (30.0%) |
| 12-18 | 315254 (27.1%) | 129434 (31.3%) | 444688 (28.2%) |
| **Sex** |  |  |  |
| Female | 554659 (47.7%) | 201959 (48.8%) | 756618 (48.0%) |
| Male | 608819 (52.3%) | 211496 (51.2%) | 820315 (52.0%) |
| **Race/ Ethnicity** |  |  |  |
| Asian Americans and<br>Pacific Islanders | 54636 (4.7%) | 19733 (4.8%) | 74369 (4.7%) |
| Hispanic | 274391 (23.6%) | 102337 (24.8%) | 376728 (23.9%) |
| Multiple | 30931 (2.7%) | 8864 (2.1%) | 39795 (2.5%) |
| Non-Hispanic Black | 195492 (16.8%) | 71564 (17.3%) | 267056 (16.9%) |
| Non-Hispanic White | 499954 (43.0%) | 171404 (41.5%) | 671358 (42.6%) |
| Other/Unknown | 108074 (9.3%) | 39553 (9.6%) | 147627 (9.4%) |
| **Site Index** |  |  |  |
| A | 89810 (7.7%) | 27544 (6.7%) | 117354 (7.4%) |
| B | 139352 (12.0%) | 53775 (13.0%) | 193127 (12.2%) |
| C | 65449 (5.6%) | 15031 (3.6%) | 80480 (5.1%) |
| D | 14396 (1.2%) | 5102 (1.2%) | 19498 (1.2%) |
| E | 39273 (3.4%) | 8230 (2.0%) | 47503 (3.0%) |
| F | 9426 (0.8%) | 4257 (1.0%) | 13683 (0.9%) |
| G | 42978 (3.7%) | 10539 (2.5%) | 53517 (3.4%) |
| H | 14386 (1.2%) | 5743 (1.4%) | 20129 (1.3%) |
| I | 28785 (2.5%) | 9193 (2.2%) | 37978 (2.4%) |
| J | 13709 (1.2%) | 5054 (1.2%) | 18763 (1.2%) |
| K | 80676 (6.9%) | 36989 (8.9%) | 117665 (7.5%) |
| L | 105681 (9.1%) | 22904 (5.5%) | 128585 (8.2%) |
| M | 43508 (3.7%) | 13741 (3.3%) | 57249 (3.6%) |
| N | 3950 (0.3%) | 1553 (0.4%) | 5503 (0.3%) |
| O | 89364 (7.7%) | 24724 (6.0%) | 114088 (7.2%) |
| P | 38952 (3.3%) | 16529 (4.0%) | 55481 (3.5%) |
| Q | 30009 (2.6%) | 8961 (2.2%) | 38970 (2.5%) |
| R | 110479 (9.5%) | 50943 (12.3%) | 161422 (10.2%) |
| S | 23140 (2.0%) | 14527 (3.5%) | 37667 (2.4%) |
| T | 3339 (0.3%) | 1383 (0.3%) | 4722 (0.3%) |
| U | 12724 (1.1%) | 23894 (5.8%) | 36618 (2.3%) |
| V | 15556 (1.3%) | 4836 (1.2%) | 20392 (1.3%) |
| W | 26780 (2.3%) | 4831 (1.2%) | 31611 (2.0%) |
| X | 38333 (3.3%) | 12499 (3.0%) | 50832 (3.2%) |
| Y | 1039 (0.1%) | 772 (0.2%) | 1811 (0.1%) |
| Z | 35262 (3.0%) | 11130 (2.7%) | 46392 (2.9%) |
| AA | 4719 (0.4%) | 2219 (0.5%) | 6938 (0.4%) |
| AB | 30205 (2.6%) | 11412 (2.8%) | 41617 (2.6%) |
| AC | 12198 (1.0%) | 5140 (1.2%) | 17338 (1.1%) |
| **Cohort Entry Period** |  |  |  |
| 03/2020 - 05/2020 | 19779 (1.7%) | 3793 (0.9%) | 23572 (1.5%) |
| 06/2020 - 08/2020 | 69928 (6.0%) | 12219 (3.0%) | 82147 (5.2%) |
| 09/2020 - 11/2020 | 98028 (8.4%) | 20709 (5.0%) | 118737 (7.5%) |
| 12/2020 - 02/2021 | 98016 (8.4%) | 37326 (9.0%) | 135342 (8.6%) |
| 03/2021 - 05/2021 | 107170 (9.2%) | 22859 (5.5%) | 130029 (8.2%) |
| 06/2021 - 08/2021 | 107358 (9.2%) | 23796 (5.8%) | 131154 (8.3%) |
| 09/2021 - 11/2021 | 162532 (14.0%) | 38181 (9.2%) | 200713 (12.7%) |
| 12/2021 - 02/2022 | 124071 (10.7%) | 123976 (30.0%) | 248047 (15.7%) |
| 03/2022 - 05/2022 | 93889 (8.1%) | 31854 (7.7%) | 125743 (8.0%) |
| 06/2022 - 08/2022 | 73250 (6.3%) | 48037 (11.6%) | 121287 (7.7%) |
| 09/2022 - 11/2022 | 113927 (9.8%) | 24082 (5.8%) | 138009 (8.8%) |
| 12/2022 - 03/2023 | 95530 (8.2%) | 26623 (6.4%) | 122153 (7.7%) |
| **Obesity** |  |  |  |
| Non-Obese | 660682 (56.8%) | 208580 (50.4%) | 869262 (55.1%) |
| Obese | 378266 (32.5%) | 163729 (39.6%) | 541995 (34.4%) |
| Unknown | 124530 (10.7%) | 41146 (10.0%) | 165676 (10.5%) |
| **PMCA Index** |  |  |  |
| 0 | 811758 (69.8%) | 289146 (69.9%) | 1100904 (69.8%) |
| 1 | 193920 (16.7%) | 69616 (16.8%) | 263536 (16.7%) |
| 2 | 157800 (13.6%) | 54693 (13.2%) | 212493 (13.5%) |
| **Number of Tests** |  |  |  |
| 0 | 856237 (73.6%) | 245524 (59.4%) | 1101761 (69.9%) |
| 1 | 193189 (16.6%) | 87927 (21.3%) | 281116 (17.8%) |
| 2 | 62803 (5.4%) | 37458 (9.1%) | 100261 (6.4%) |
| 2+ | 51249 (4.4%) | 42546 (10.3%) | 93795 (5.9%) |
| **Number of Drugs** |  |  |  |
| 0 | 278662 (24.0%) | 80867 (19.6%) | 359529 (22.8%) |
| 1 | 137488 (11.8%) | 45771 (11.1%) | 183259 (11.6%) |
| 2 | 111410 (9.6%) | 40194 (9.7%) | 151604 (9.6%) |
| 2+ | 635918 (54.7%) | 246623 (59.6%) | 882541 (56.0%) |

The incidence of individual and composite GI conditions was higher among COVID-19 positive patients than among COVID-19 negative patients, which were summarized in **Table 2**. We showed the aRR for each GI symptom and disorder and composite outcomes during the post-acute phase or the chronic phase in **Figure 2**. The SMDs of all baseline covariates were balanced (less than 0.1) shown in **Section S1** of the Supplement. We observed significantly elevated risks across various GI outcomes when comparing COVID-19 positive patients to their COVID-19 negative counterparts. Specifically, the risk of experiencing abdominal pain was higher in the COVID-19 positive group (2.54%) compared to the COVID-19 negative group (2.06%), with an aRR of 1.14 (95% CI, 1.11-1.17). Bloating was more prevalent in the COVID-19 group (0.28% vs. 0.23%), with an aRR of 1.27 (95% CI, 1.18-1.37). Constipation also showed an increased incidence (2.94% vs. 2.42%), with an aRR of 1.20 (95% CI, 1.17-1.23). Diarrhea was reported more frequently (2.30% vs. 1.57%), with an aRR of 1.40 (95% CI, 1.36-1.43). Nausea and vomiting were similarly elevated, with nausea at 0.81% versus 0.56% (aRR, 1.27; 95% CI, 1.21-1.33) and vomiting at 2.98% versus 2.29% (aRR, 1.33; 95% CI, 1.30-1.36). GERD also exhibited a higher prevalence, 1.15% versus 1.00%, with an aRR of 1.19 (95% CI, 1.15-1.24). However, the risk of IBS did not have statistical significance after adjusting for confounding variables (0.10% vs. 0.11%; aRR, 0.91; 95% CI, 0.81-1.02). Additionally, the COVID-19 positive cohort exhibited increased risks for a composite of any of these GI signs or symptoms (8.24% vs. 6.45%; aRR, 1.26; 95% CI: 1.24-1.28), any of these GI disorders (1.27% vs. 1.12%; aRR, 1.19; 95% CI, 1.15-1.24), and any visits related to GI (8.64% vs. 6.85%; aRR, 1.19; 95% CI, 1.15-1.24).

**Figure 2.**
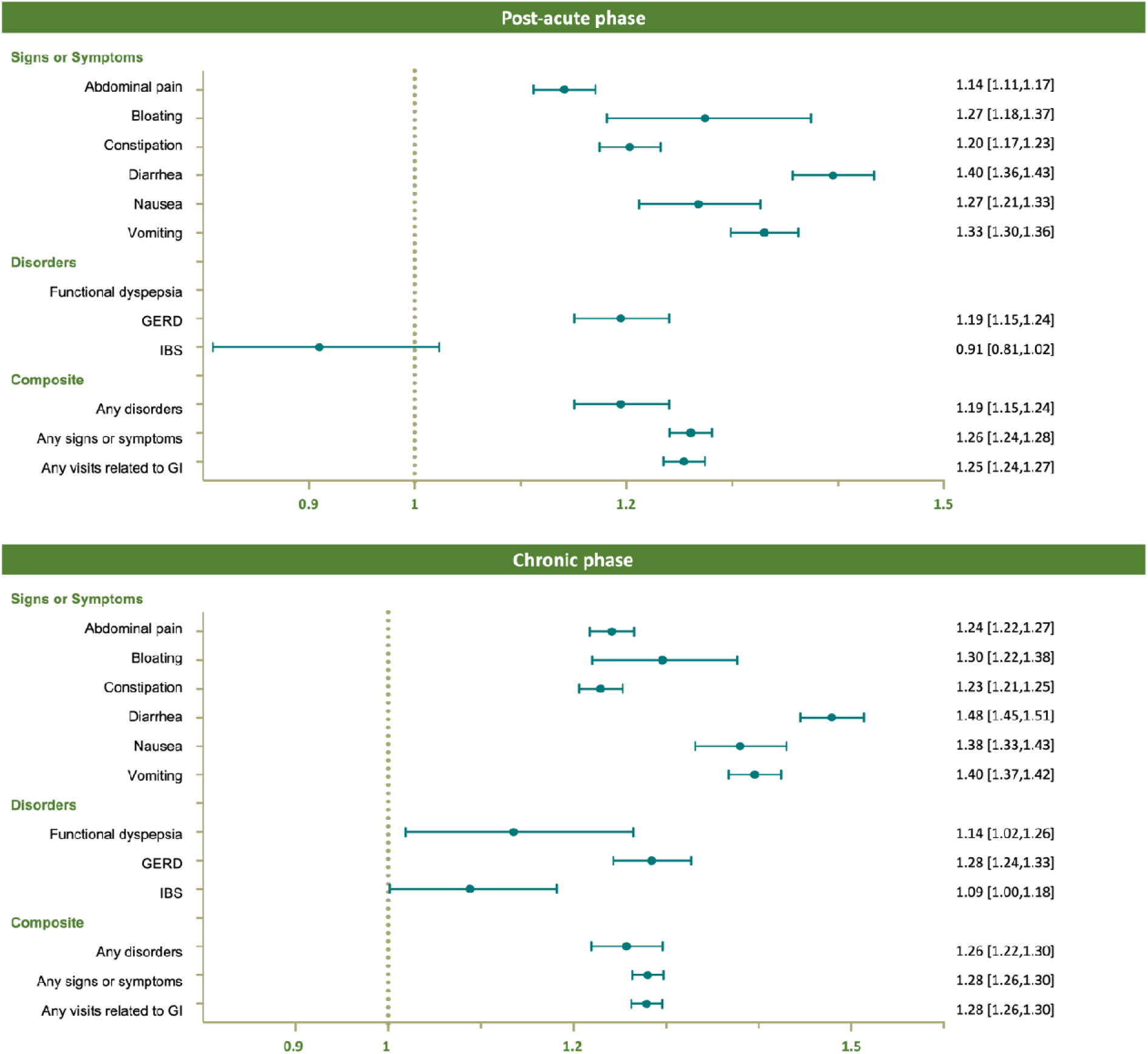
Adjusted risk ratio for the post-acute phase (28 days to 179 days after SARS-CoV-2 infection) and the chronic phase (180 days to 729 days after SARS-CoV-2 infection) comparing COVID-19 positive patients to COVID-19 negative patients.

**Table 2.**
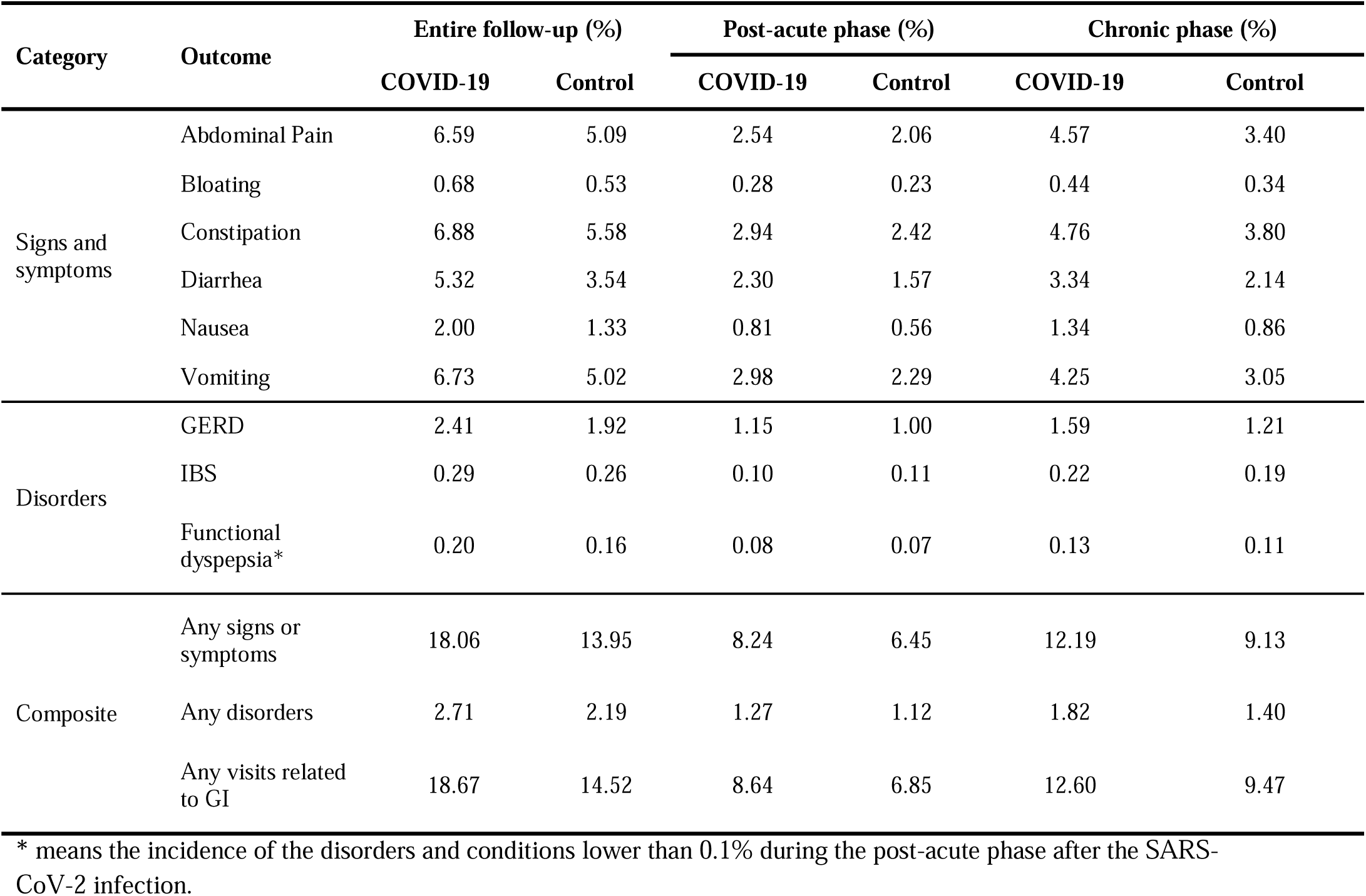
Incidence table in % for post-acute phase or chronic phase of COVID-19 infection.

| Category | Outcome | Entire follow-up (%) |  | Post-acute phase (%) |  | Chronic phase (%) |  |
| --- | --- | --- | --- | --- | --- | --- | --- |
|  |  | COVID-19 | Control | COVID-19 | Control | COVID-19 | Control |
| Signs and symptoms | Abdominal Pain | 6.59 | 5.09 | 2.54 | 2.06 | 4.57 | 3.40 |
|  | Bloating | 0.68 | 0.53 | 0.28 | 0.23 | 0.44 | 0.34 |
|  | Constipation | 6.88 | 5.58 | 2.94 | 2.42 | 4.76 | 3.80 |
|  | Diarrhea | 5.32 | 3.54 | 2.30 | 1.57 | 3.34 | 2.14 |
|  | Nausea | 2.00 | 1.33 | 0.81 | 0.56 | 1.34 | 0.86 |
|  | Vomiting | 6.73 | 5.02 | 2.98 | 2.29 | 4.25 | 3.05 |
| Disorders | GERD | 2.41 | 1.92 | 1.15 | 1.00 | 1.59 | 1.21 |
|  | IBS | 0.29 | 0.26 | 0.10 | 0.11 | 0.22 | 0.19 |
|  | Functional dyspepsia* | 0.20 | 0.16 | 0.08 | 0.07 | 0.13 | 0.11 |
| Composite | Any signs or symptoms | 18.06 | 13.95 | 8.24 | 6.45 | 12.19 | 9.13 |
|  | Any disorders | 2.71 | 2.19 | 1.27 | 1.12 | 1.82 | 1.40 |
|  | Any visits related to GI | 18.67 | 14.52 | 8.64 | 6.85 | 12.60 | 9.47 |

\* means the incidence of the disorders and conditions lower than 0.1% during the post-acute phase after the SARS-CoV-2 infection.

In the chronic phase shown in **Figure 2**, COVID-19 positive patients exhibited elevated risks of various GI symptoms and symptoms and disorders compared to the COVID-19 negative cohort. Specifically, the heightened risk persisted for a composite of any of these GI signs or symptoms (12.19% vs. 9.13%; aRR, 1.26; 95% CI: 1.22-1.30), any of these GI disorders (1.82% vs. 1.40%; aRR, 1.28; 95% CI, 1.26-1.30), and any visits related to the GI (12.60% vs. 9.47%; aRR, 1.28; 95% CI, 1.26-1.30). These findings highlighted the persistent higher health risks among individuals with COVID-19 across a range of GI outcomes for the chronic phase. Moreover, when compared to the post-acute phase, the aRR for each GI outcome was higher in the chronic phase, indicating a prolonged impact of the infection on GI health.

We presented the results of the subgroup analysis for composite outcomes in **Figure 3**, categorized by age, race/ethnicity, sex, cohort entry period, medical history of cardiovascular disease, diabetes, obesity status, the severity of the acute phase of COVID-19, and hospitalization status during the acute phase of COVID-19. Our findings revealed that children under the age of five were at the highest risk for GI disorders or symptoms compared to other age groups. Detailed results for the specific GI symptoms and disorders by age are shown in **Tables S1-S3**. The aRR did not vary across different racial/ethnic groups, as presented in **Tables S4-S7**. Interestingly, obese patients had a lower risk of GI symptoms or disorders compared to non-obese patients (**Tables S8-S9**). Male patients exhibited a higher risk of GI disorders or symptoms during the post-acute phase compared to female patients (**Tables S10-S11**). No significant differences in the risk of GI symptoms and disorders were found between patients with and without diabetes (**Tables S12-S13**) or between those with and without cardiovascular disease (**Tables S14-S15**). However, the aRR of GI-related visits increased progressively from the non-hospitalized group to the hospitalized group, and finally to the ICU admission group (**Tables S16-S19**). Furthermore, the aRR of GI-related visits rose with the severity of the acute phase of COVID-19, escalating from the mild to the moderate and severe groups (**Tables S20-S22**). Compared with the Omicron or Delta period, the risk of GI symptoms during the Pre-Delta period was increased (**Tables S23-S25**). The SMD for different subgroup analyses was less than 0.1 and the empirical clinical equipoises^40^ lied within the reasonable range (30%-70%) shown in **Sections S1-S10**. We conducted negative control outcome experiments as a sensitivity analysis in **Section S11**, which indicated a slight systematic error, evidenced by a minor shift in point estimates with wider CIs.

**Figure 3.**
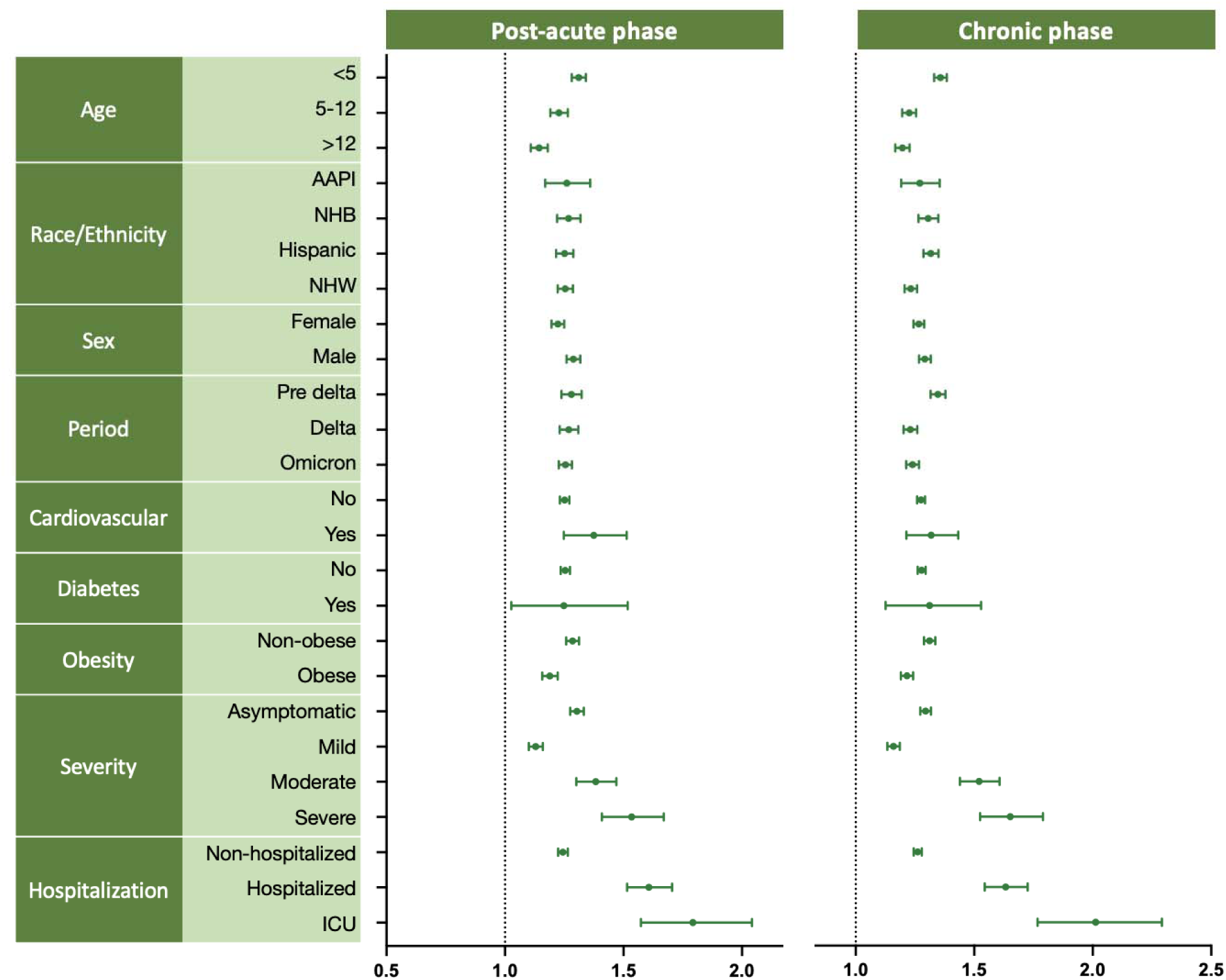
A subgroup analysis for the adjusted risk ratios of GI visits between COVID-19 positive patients an COVID-19 negative patients. It details the adjusted risk ratios for two distinct phases: the post-acute phase, spanning from 28 to 179 days after SARS-CoV-2 infection, and the chronic phase, ranging from 180 to 729 days post-infection.

## Discussion

### Principal Findings

In this population-based study of 1,576,933 US patients under eighteen, we found a significant increase in the risk of various GI outcomes associated with COVID-19, including abdominal pain, bloating, constipation, diarrhea, nausea, vomiting, and GERD. COVID-19 was also linked to a higher risk of experiencing at least one GI symptom, disorder, or GI-related visit during both the post-acute and chronic phases. Our study is the largest study reported for children and adolescents that assesses the risks of COVID-19 infection for GI symptoms and diseases in the US.

### Comparison with Other Studies

Our findings align with prior research indicating that COVID-19 elevates the risk of GI symptoms such as nausea, diarrhea, and disorders like functional dyspepsia^41–46^. Many of these earlier studies were limited by insufficient sample sizes for precise estimations and restricted their risk assessments to specific geographic regions and time frames^41–44^. Our results are also consistent with a study that utilized the US Department of Veterans Affairs national health care databases to examine increased gastrointestinal symptom or disorder risks in the post-acute phase following infection^21^. Similar to our findings, this study observed a significantly higher risk of GI symptoms and disorders among SARS-CoV-2-infected individuals, including abdominal pain, bloating, constipation, diarrhea, GERD, and vomiting. Our study uniquely focuses on children and adolescents, extending the evaluation of risk into the chronic phase beyond the post-acute period.

### Interpretation

One potential biological mechanism underlying this association is the high expression of angiotensin-converting enzyme 2 on the brush border of the small intestinal mucosa.^47^ Furthermore, SARS-CoV-2 infection has been shown to influence the gut microbiome^48^, and this impact potentially persists beyond the post-acute phase^49^. Additionally, evidence from studies on prolonged viral fecal shedding^50^ and the persistence of the virus in the GI tract^51^ lends further support to the connection we have observed. A more profound understanding of the biological mechanisms will contribute to the development of targeted and effective interventions, ultimately improving outcomes for those affected by long-term GI disorders.

### Strengths and Limitations

An outstanding feature of this study is its utilization of population-based databases within RECOVER. This large program facilitated the precise identification of comprehensive COVID-19 infection records and visits associated with GI symptoms and disorders throughout the entire study period in the US, thereby mitigating the potential for selection bias. Furthermore, it provided detailed and accurate information on infection status, reducing the likelihood of exposure misclassification bias. Notably, our investigation extended into the chronic phase, with a follow-up period of up to two years, representing one of the longest follow-up durations reported for children and adolescents exposed to COVID-19 infection in the US.

Our study has some limitations. Firstly, while we successfully achieved a balanced distribution of baseline covariates through matching, the propensity scores were constructed using only the variables accessible within our study databases. Nevertheless, the confounder list lays a solid foundation for our propensity score model, enhancing the reliability and validity of our analyses.

Besides, it is important to highlight that our findings remained consistent and robust through sensitivity analyses aimed at addressing residue confounding by negative control outcomes. Moreover, despite the large sample size, our study might have ruled out small associations for rare events, such as functional dyspepsia in the post-acute phase. We ruled out functional dyspepsia in post-acute phase analysis due to its rare occurrence (<0.1%), but we observed an increased aRR in functional dyspepsia during the chronic phase. Furthermore, while our findings indicate a moderate reduction in the aRR for IBS during the post-acute phase, it is important to note that despite abdominal pain being a primary symptom of pediatric IBS, the Rome IV criteria^52^ require a minimum of six months from symptom onset for a diagnosis of IBS. This timeframe typically extends beyond the post-acute period, complicating the diagnosis within that phase. Additionally, the misclassification of documented SARS-CoV-2 infection can be a problem within EHR. However, recent studies^53^ provided a novel statistical method to account for the misclassified documented infection with prior knowledge about the prespecified range of misclassification rates given the patients get infected with SARS-CoV-2 infection or diagnosed with COVID-19. Implementing this statistical approach helps to mitigate some of the biases introduced by EHR misclassifications and strengthens the reliability of our findings. However, future studies will need to continue refining these techniques and exploring additional methods to further reduce the impact of misclassification on GI outcomes. This ongoing research is pivotal not only for improving the clinical management of children with persistent GI symptoms and disorders, but also for informing public health strategies aimed at preventing long-term consequences of SARS-CoV-2 infection.

### Disclaimer

This content is solely the responsibility of the authors and does not necessarily represent the official views of the RECOVER Initiative, the NIH, or other funders.

## Data Availability

The data can be shared with the request to the RECOVER initiative.

## Author Contributions

Authorship was determined using *ICMJE* recommendations. DZ, RS, TZ, and YL designed methods and experiments; CF provided the datasets for data analysis; DZ and YL conducted data analysis; all coauthors interpreted the results and provided instructive comments; DZ, RS, TZ, and YL drafted the main manuscript. All coauthors provided critical edits to the early draft and approved the final version of the manuscript. The corresponding author attests that all listed authors meet authorship criteria and that no others meeting the criteria have been omitted.

## Funding

This research was funded in part by the National Institutes of Health (NIH) Agreement OTA OT2HL161847-01 as part of the Researching COVID to Enhance Recovery (RECOVER) Research Initiative.

## Potential Conflicts of Interest

Dr. Becich reports PCORnet funding as MPI for the PaTH Clinical Research Network. All other authors have no conflicts of interest to report.

## Ethical Approval

This study was approved by the University of Pennsylvania’s institutional review board (No. 851604), with a waiver of informed consent.

## Data Availability

The data can be shared with the request to the RECOVER initiative.

## Acknowledgments

This study is part of the NIH Researching COVID to Enhance Recovery (RECOVER) Initiative, which seeks to understand, treat, and prevent the post-acute sequelae of SARS-CoV-2 infection (PASC). For more information on RECOVER, visit https://recovercovid.org/. We would like to thank the National Community Engagement Group (NCEG), all patients, caregivers, and community Representatives, and all the participants enrolled in the RECOVER Initiative. A special thanks to patient representatives Megan Carmilani and Etienne Carignan for their helpful comments and suggestions.

## Notes

### Author Declarations

This study was approved by the University of Pennsylvania institutional review board (No. 851604) with a waiver of informed consent.

### Summary of Updates

The section on Results was updated to clarify Figure 3. Abstract revised.

